# Systematic review and meta-analysis of child and adolescent mental health in South Africa

**DOI:** 10.1101/2025.09.09.25335400

**Authors:** Jason Bantjes, Dan Jenkins, Carrie Brooke-Sumner, Lauro Estivalete Marchionatti, Noluthando Mpisane, Meghan Mosalisa, Yeukai Chideya, Tenielle C. Pengelly, Dan J. Stein, Soraya Seedat, Mark Tomlinson, Sarah Skeen, Anusha Lachman, Inge Petersen, Bonga Chiliza, Rene Nassen, Saeedda Paruk, Merryne Young, Xanthe Hunt, Nuhaa Holland, Shirley Reynolds, Eleanor Chatburn, Elelwani Charity Mamathuba, Zeina Mneimneh, Giovanni A Salum

**Affiliations:** Mental Health, Alcohol, Substance use, and Tobacco Research Unit, South African Medical Research; Department of Psychiatry and Mental Health, University of Cape Town; Stavros Niarchos Foundation Global Center for Child and Adolescent Mental Health, Child Mind Institute, New York, NY, USA; Stellenbosch University, Cape Town, South Africa; Mental health, Alcohol, Substance use, and Tobacco Research Unit, South African Medical Research Council, South Africa; Department of Psychiatry, Universidade Federal do Rio Grande do Sul, Porto Alegre, RS, Brazi; tavros Niarchos Foundation Global Center for Child and Adolescent Mental Health, Child Mind Institute, New York, NY, USA; Mental health, Alcohol, Substance use, and Tobacco Research Unit, South African Medical Research; Institute for Life Course Health Research, Department of Global Health, Stellenbosch; Mental health, Alcohol, Substance use, and Tobacco Research Unit, South African Medical Research Council; Institute for Life Course Health Research, Department of Global Health, Stellenbosch University, Cape Town, South Africa; Department of Psychiatry and Mental Health, University of Cape Town, South Africa; Department of Psychiatry, Stellenbosch University; Department of Psychiatry, Stellenbosch University, Tygerberg Campus, Parow 7550, South Africa; Centre for Rural Health, School of Nursing and Public Health, University of Kwazulu-Natal, Howard College; Department of Psychiatry, University of KwaZulu-Natal, Durban, South Africa; Africa Health Research Institute, South Africa; Global Programs, Child Mind Institute, New York, NY, USA; Department of Psychiatry, Universidade Federal do Rio Grande do Sul & Hospital de Clínicas de Porto Alegre, Porto Alegre, Brazil

**Keywords:** Mental health, mental disorders, suicide, children, adolescents, South Africa, systematic review, meta-analysis

## Abstract

**Introduction:** Reliable epidemiological data are crucial to make evidence-based decisions about youth mental health. Yet little is known about the epidemiology of child and adolescent mental health in sub-Saharan Africa. We conducted a systematic review and meta-analysis on the prevalence of mental disorders, suicidal behaviors, mental well-being and mental health awareness/literacy among children and adolescents in South Africa (SA).

**Methods:** We searched PubMed, PsycINFO, Web of Science, Scielo.org, and Google Scholar from their inception to 19th February 2025. We performed random effects meta-analysis for all disorders that had 5 or more prevalence estimates. Meta-regressions were used to investigate factors associated with prevalence estimates.

**Results:** We screened 12,768 records and identified 40 studies with 56 prevalence estimates for mental disorders and 30 prevalence estimates for suicidality. Across all studies on mental disorders, the pooled prevalence for all disorders was 8.53% [6.1; 11.9] (k=56, N=39,962), with significant heterogeneity (I²=99.0%, Q (55) = 5467.0, p<.001). Pooled prevalence estimates for depressive disorders, anxiety disorders, PTSD and behavioural disorders were 10.1% [4.9; 19.9], 6.7% [3.4; 12.8], 17.6% [8.5; 33.1], 3.9% [1.8; 8.5], respectively. All other disorders had 5 or fewer prevalence estimates. Pooled prevalence estimates for suicidal ideation, plan and attempt were 12.0% [7.8; 18.0] (k=10, N=41489), 11.8% [7.7; 17.6] (k=8, N=39,928), and 10.3% [6.2; 16.6] (k=9, N=40,294), respectively. No papers reported mental well-being, quality of life, mental health literacy, mental health awareness, or cognitive impairment. It is not possible to reliably assess the mental health of SA’s youth due to the small number of studies, narrow focus on few disorders and heterogeneity.

**Conclusion:** There is clear need for a reliable national survey of child and adolescent mental health in SA, using well validated instruments that can assess a wide range of disorders and mental well-being among a representative sample of young people.

## INTRODUCTION

The mental health of children and adolescents is an important public health priority[1,2], with the global burden of mental disorders among young people increasing significantly in the past 30 years[3]. Most mental disorders have their onset early in life[4,5] and, left untreated, have a long-lasting deleterious impact on development. Nonetheless, the mental health needs of children and adolescents are often neglected, especially in low- and middle-income countries[6]. Reliable epidemiological data are crucial to address the burden of mental health problems in young people, plan interventions, and set priorities and policies[7,8], yet data on mental health remain sparse[9], with no country in sub-Saharan Africa having data for more than 2% of the population of children and adolescents[10]. Indeed, very little is known about the epidemiology of child and adolescent mental health in sub-Saharan Africa[11].

South Africa (SA) is an upper middle-income country with a population of approximately 62 million people, 34% of which are under 18 years of age. The country has a history of colonisation and racial segregation under Apartheid, which has created persistent problems with inequality and poverty. Although the first democratic elections took place in 1994, the country continues to struggle to address structural issues which contribute to inequality, a high burden of infectious diseases, high rates of unemployment, crime, violence, and hazardous substance use. As many as 53% of children live in poverty[12], with approximately two thirds of children exposed to community violence and more than half exposed to violence in their own homes[13]. Further, 39% of girls report having undergone some form of sexual violence (e.g., unwanted touching, forced sex, or exploitation) before the age of 18 years[14]. These conditions create vulnerability for serious and persistent mental health problems among young people. Yet, it remains unclear what the burden of mental health problems is amongst SA youth. Reliable and comprehensive data are urgently needed to better understand the burden of disease and mental healthcare needs of SA’s young people. Without these data it is not possible to advocate for appropriate evidence-based policies or facilitate equitable preventive healthcare services for SA’s child and adolescent population[15].

Our aim is to synthesize available evidence on prevalence of mental disorders. We conducted a systematic review and meta-analysis of the prevalence of mental disorders, suicidal thoughts and behaviours, and mental well-being among children and adolescents (i.e. ≤18 years-old) in SA. This work is part of a larger Global Landscaping initiative by the Stavros Niarchos Foundation (SNF) *Global Center for Child and Adolescent Mental Health* at the *Child Mind Institute* (CMI), who have conducted similar systematic reviews in Greece[16] and Brazil[17]. The SA component of this landscaping is being conducted in partnership with the *South African Medical Research Council* (SAMRC) and local experts, as a first step towards compiling a comprehensive research agenda and roadmap to guide policy and practice to promote child and adolescent mental health in the country. In addition to this review, reviews on substance use among young people, the translation and validation of psychometric instruments for assessing mental health, and psychosocial interventions to promote child and adolescent mental health in SA are underway.

## METHODS

The approach we adopted was informed by a highly cited meta-analysis on child and adolescent mental health[18]. The review protocol was registered on PROSPERO[19].

### Search Strategy

To identify studies for this review and the subsequent planned reviews described above, we conducted one comprehensive, multi-step procedure to search PubMed, PsycINFO, Web of Science, and Scielo.org (SA Collection), without restrictions of language or date, using English search terms related to children and adolescents, SA, mental health, mental disorders, and suicide (see supplementary materials Table S1). We also conducted a complementary search on Google Scholar using broad search terms (i.e., “mental” AND (“child” OR “adolescent” AND “South Africa”), to identify any records that may have been missed in the other databases. Two researchers screened the Google Scholar search results for new inclusions, until reaching a point of scanning 50 consecutive records without identifying any new records. Backward citation searching of reference lists of systematic reviews identified in these searches was also conducted to identify additional potentially relevant studies. The initial searches were done on 20^th^ March 2024 and then updated on 19^th^ February 2025.

### Data management

Deduplication was done using the software EPPI-Reviewer 4.0, with the calibration index for detection set to 0.85, which has been shown to yield a low rate of false positives [20]. Studies were then uploaded to Rayyan for data management[21] and an additional step of deduplication was performed.

### Screening procedures

Four researchers (NM, YC, NH and MM) worked independently reviewing all records by title and abstract to identify potential inclusions. All records were reviewed independently by two researchers with disputes resolved through discussion and consultation with a third reviewer (CB). Prior to full text screening and data extraction, the members of the team met to collectively review a random subset of 10 studies to ensure shared understanding of inclusion/exclusion criteria. Database searches identified 12,761 records for review after removing duplicates.

### Inclusion and exclusion criteria

To be included, studies needed to meet all the following inclusion criteria:

1. Report original data for SA collected via either epidemiological registers or surveys with community-based and/or school-based samples.
2. Report on a sample of children and/or adolescents ≤18 years-old or with mean age ≤18 years if older individuals were included.
3. Data collected from self-report and/or informants (i.e. parents, caregivers, teachers, etc.).
4. Results contained prevalence estimates for any of the following domains of child and adolescent mental health:

- *Mental disorders* (as defined by ICD or DSM) and/or suicidal behaviours and/or self-harm assessed in any of the following ways: diagnosis determined by a clinical interview; diagnosis determined by cut-off on a validated or standardized screening instrument / structured questionnaire; symptom severity (i.e. mild/moderate/severe) assessed via screening instruments.
- *Mental well-being* and/or *quality of life* (e.g. sleep, subjective mental well-being, self-esteem, resilience, belonging level of impairment/functioning) assessed via a validated or standardized instrument / structured questionnaire.
- *Mental health literacy* and/or *mental health awareness*.
- Measures of *cognitive development* (e.g. reading age, spelling problems, school readiness, etc.).
5. Findings published in any of the following formats: journal articles, editorial letters if original data reported, book chapters if original data reported, or reports of national surveys.

Studies were excluded if they met *an*y of the following criteria:

1. Samples not drawn from the general population (i.e. samples entirely from populations or groups with very specific characteristics that made them non-representative of the general population, such as survivors of traumatic events, juvenile offenders, HIV positive children, clinical samples). This is not to say that studies which included some youth with any of these conditions were excluded, it only means that studies that reported exclusively on youth with these conditions were excluded because they focus on special populations rather than the general population.
2. Studies that reported *any* of the following outcomes (unless they also included prevalence data on the mental health outcomes for children/adolescents as listed above):

- motor and/or language development (e.g. verbal fluency, phonetic awareness, fine motor control).
- maltreatment, adverse childhood events, and/or exposure to violence.
- data on the mental health of caregivers or childcare professionals (e.g. maternal depression, teacher burnout).
3. Studies which only reported the absolute number of cases without rates (i.e. counts with no denominators).

In cases where secondary data from a national survey were reported, we excluded the publication and substituted it with the original report of the national survey so that we only included primary data. If the same data were reported in more than one publication, we included the earliest publication and excluded any subsequent later publications. We excluded studies that assessed prevalence rates during the COVID period of social distancing (i.e. 2020/21) and studies reporting modelled data (e.g.,Global Burden of Disease data).

### Data extraction

Data was extracted by reviewers (NM, YC, NH and MM) using a data extraction form programmed in RedCap. The data extraction form was pilot tested with a subset of 10 articles before it was finalised. Detailed data were extracted about the study year, study design, study setting, methods, sample characteristics, criteria for determining caseness, psychometric instruments used, and results. Our decision about what data to extract was informed by a prior large systematic review and meta-analysis in this field[22]. All extracted data used in the final tables were checked for accuracy and completeness by a second reviewer (TCP) who referred to the original sources.

### Quality assessment

We used the *Joanna Briggs Institute* (JBI) critical appraisal tool for prevalence studies[23] to assess the quality of included studies and assess risk of bias. These quality assessments were conducted by one author (MM) and then checked by a second author (TCP).

### Data analysis

Data were synthesized and summarized into tables and figures using Excel. In studies where the prevalence estimate was obtained using a screening instrument (i.e. reporting apparent prevalence rates) a Bayesian estimation of true prevalence was calculated using the Beta-expert method[24,25]. These calculations were done using the Shiny App[26]. This approach uses Bayes’ Theorem to adjust the apparent prevalence estimates by accounting for likely false positives and false negatives using the sensitivity and specificity of the screening instrument[27]. To perform these adjustments, we used sensitivity and specificity values from validation studies conducted in SA with children and adolescents. Where there were no SA validation studies of the instrument, pooled sensitivity and specificity values from systematic reviews of validation studies with children and adolescents were used. In cases where there were no systematic reviews, we assumed with a 90% confidence interval that the sensitivity and specificity were between 0.7 and 0.9. See supplementary table S2 for details of the sensitivity and specificity values we used and the relevant sources.

We conducted a meta-analysis for all disorders that had 5 or more included studies, using logit-transformed true prevalence estimates for studies that used screening instruments and the reported prevalence for studies that used diagnostic interviews to identify cases. For the meta-analysis we pooled all studies irrespective of the recall period, because there were so few studies.

Random effects meta regression analysis was used to investigate sources of heterogeneity using the study characteristics as covariates. In these models the study characteristics we considered were: year of publication; data source (i.e. community-based survey, school-based survey or register); geographic region where data were collected; mean age of the sample, proportion of the sample identified as female; sampling procedures (probability vs convenience), whether cases were identified using symptom severity assessed with a screening instrument or diagnostic criteria / structured clinical interview; if symptoms were identified by self-report or observation, and the recall period (1-week, 2-week, 1-month, 6-months, 12-months, lifetime). We did not include population groups as a covariate because studies typically did not report these data in a meaningful way. Each of these covariates were entered individually in univariate analysis and then together in a multivariate model. For all tests of statistical analysis, Alpha was set to 0.05. The meta-analysis and meta-regression were performed by DJ using R version 4.5.0[28] and the R metafor package version 4.8.0[29]. All data files and R code are included as supplementary materials.

The *Preferred Reporting Items for Systematic Reviews and Meta-Analysis* (PRISMA) statement[30] and the ROBIS tool for assessing risk of bias in systematic reviews[31] were used to inform the reporting of findings.

### Ethics statement

This study does not involve human participants.

### Patient and public involvement

Patients and the public were not involved in this study as it is a secondary analysis of existing data. Results will be disseminated to stakeholders through networks of the SNF *Global Center for Child and Adolescent Mental Health* at CMI, and SAMRC.

## RESULTS

The screening process is summarised in the PRISMA diagram in Figure 1. A total of 12,788 records were screened by title and abstract, of which 305 were selected for full-text review. Following full-text review, we identified 40 studies that met inclusion criteria. Of the 40 included papers, 29 reported prevalence estimates for mental disorders, 10 reported prevalence estimates for suicidal thoughts and behaviours, and 1 reported prevalence estimates for both mental disorders and suicidality. There were no papers reporting on mental well-being, quality of life (e.g. sleep, subjective mental well-being, self-esteem, resilience, belonging, etc.), mental health literacy, mental health awareness, or cognitive development. Full details of all included studies are provided as supplementary materials (see Supplementary Tables S2 and S3).

**Figure 1:**
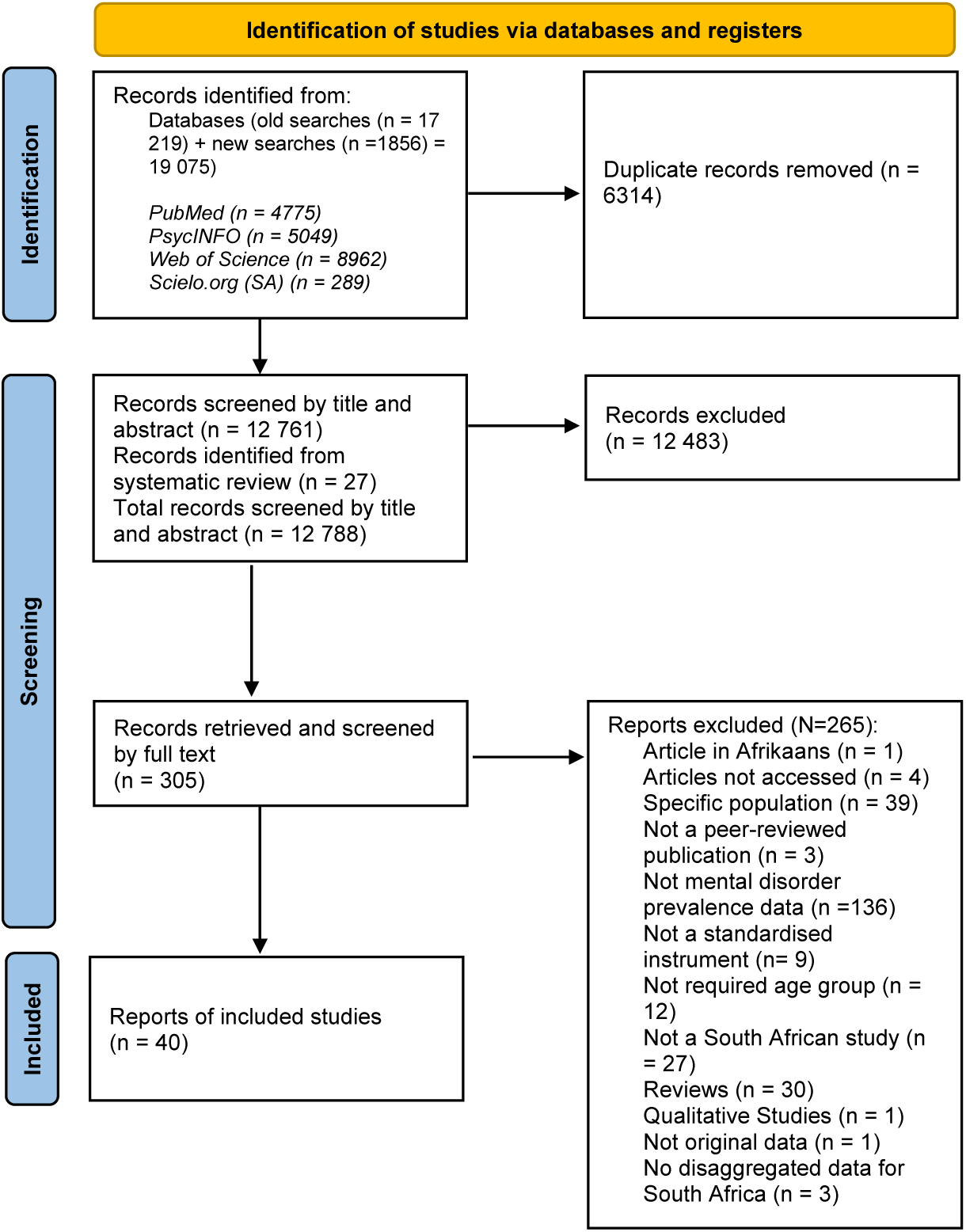
PRISMA flow diagram

Across all included studies (N=40) the pooled sample size was 70,703 (excluding one study[32] which used registry data to estimate suicide rates nationally and the total sample size was not clear). The mean age of samples was 14.3 years, for studies that reported mean age. On average, the proportion of females in the samples was 54.1%.

There was an uneven geographic distribution of studies across the 9 provinces, with only 6 studies reporting data collected nationally. Most studies reported data from the Western Cape (WC, n=21), KwaZulu-Natal (KZN, n=13) and Gauteng (GP, n=10). The heat map in Figure 2 illustrates the pooled sample size by province, showing clearly that most data come from WC (n=16,209), KZN (n=9,346) and GP (n=9,265). Supplementary Table S4 shows the number of studies (excluding one study[32] which used registry data to estimate suicide rates nationally) and pooled sample size by province relative to the size of the population of children (<18 years) in each province[33]. Although only 10% of children live in the Western Cape[33], 22.9% of the data on mental health comes from this province.

**Figure 2:**
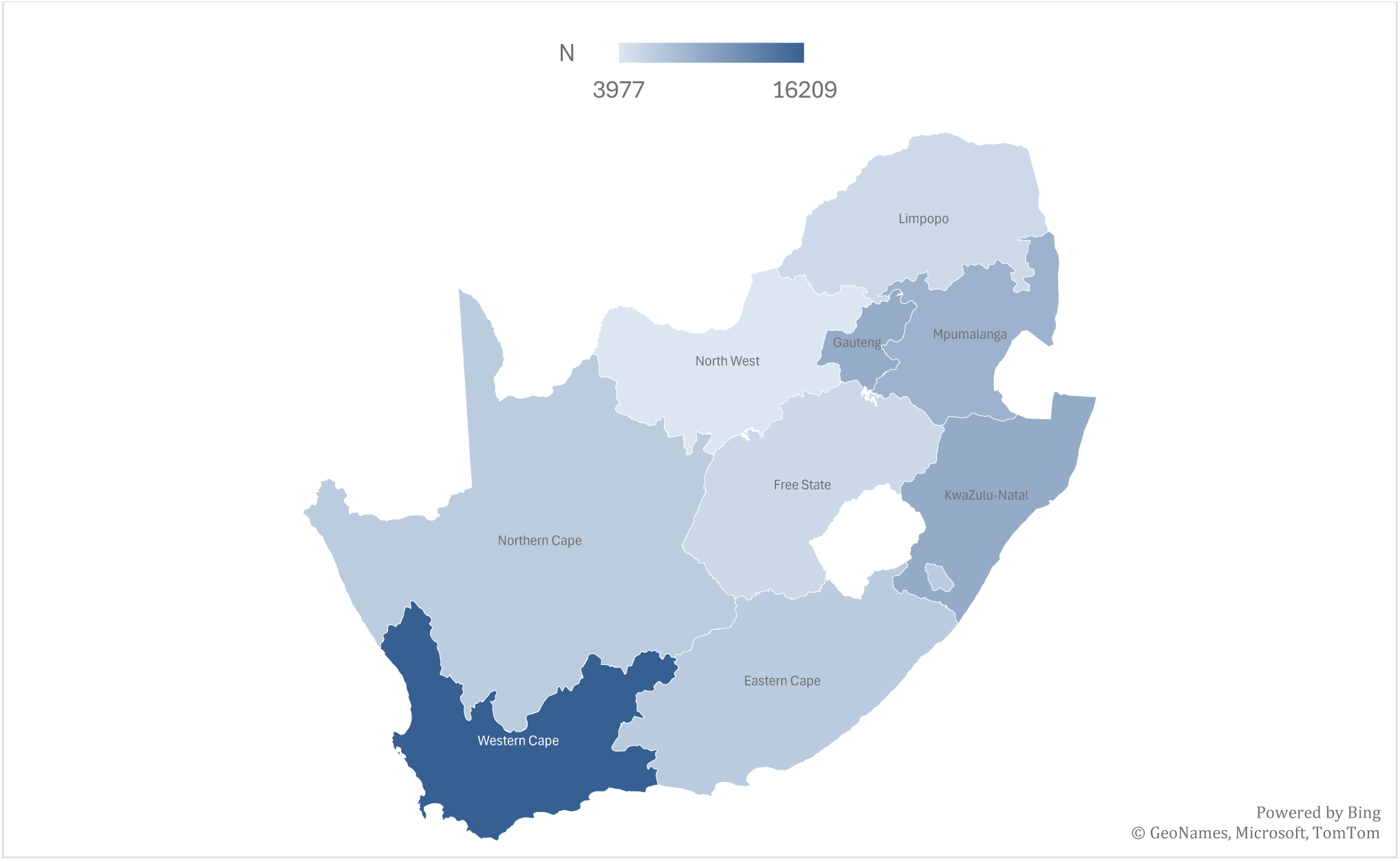
Heat map showing the pooled sample size across all included studies by province in South Africa

### Mental disorders

As shown in Figure 3, 56 prevalence estimates were reported for mental disorders, the most common of which were depressive disorders (n=15), PTSD (n=9), anxiety disorders (n=8), disruptive behaviour disorders (n=6), and ADHD (n=4). Across the 56 prevalence estimates, the majority of were derived from school-based samples (n=34), self-report data (n=28), cross-sectional studies (n=55), probability samples (n=32), and screening instruments rather than diagnostic interviews (n=36). The most common screening instruments were the *Child Behaviour Checklist* (n=6), *Centre for Epidemiologic Studies Depression Scale* (n=6), *Strengths and Difficulties Questionnaire* (n=6). The most common diagnostic interviews were the *National Institute of Mental Health’s Diagnostic Interview Schedule for Children* (n=6) and the *Diagnostic Interview Schedule for Child* (n=3).

**Figure 3:**
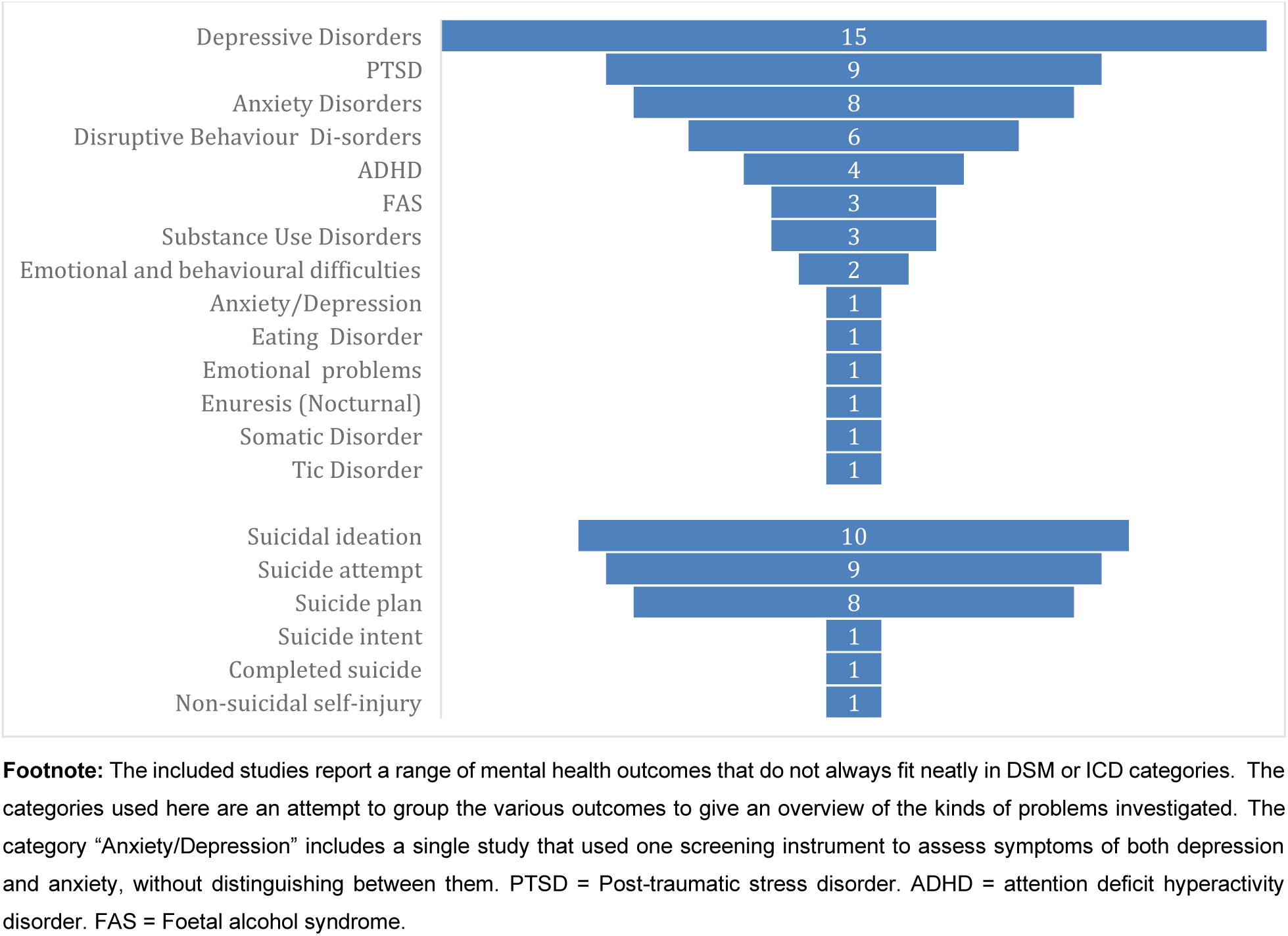
Funnel diagram showing the number of prevalence estimates for mental health conditions by disorder (n=56) and suicidal thoughts and behaviours s (n=30)

Meta-analyses were conducted on the 4 groups of disorders which had 5 or more prevalence estimates (see Figure 4). The pooled prevalence for depressive disorders of 10.1% [4.9; 19.9], (k=15, N=12,797) with significant and substantial heterogeneity among the studies (**I²** = 99.28%, Q(14)= 1949.9, p<0.0001). For PTSD, the pooled prevalence was 17.6% [8.5; 33.1] (k=9, N=5,749) with significant heterogeneity (I² =99.29%, Q(8)=360.0, p<0.0001). Anxiety disorders had a pooled prevalence of 6.7% [3.4; 12.8] (k=8, N=4,061) with included studies also showing significant heterogeneity (I²=96.85%, Q(7)=286.2, p<0.0001). Finally, the pooled prevalence for disruptive behaviour disorders was 3.9% [1.8; 8.5] (k=6, N=3,461) with significant heterogeneity among the included studies (I²=95.35%, Q(5)=62.6, p<0.0001). The pooled prevalence across all disorders was 8.53% [6.1; 11.9] with significant heterogeneity across the prevalence estimates (I²=99.0%, Q(55)= 5467.0, p<.001).

**Figure 4:**
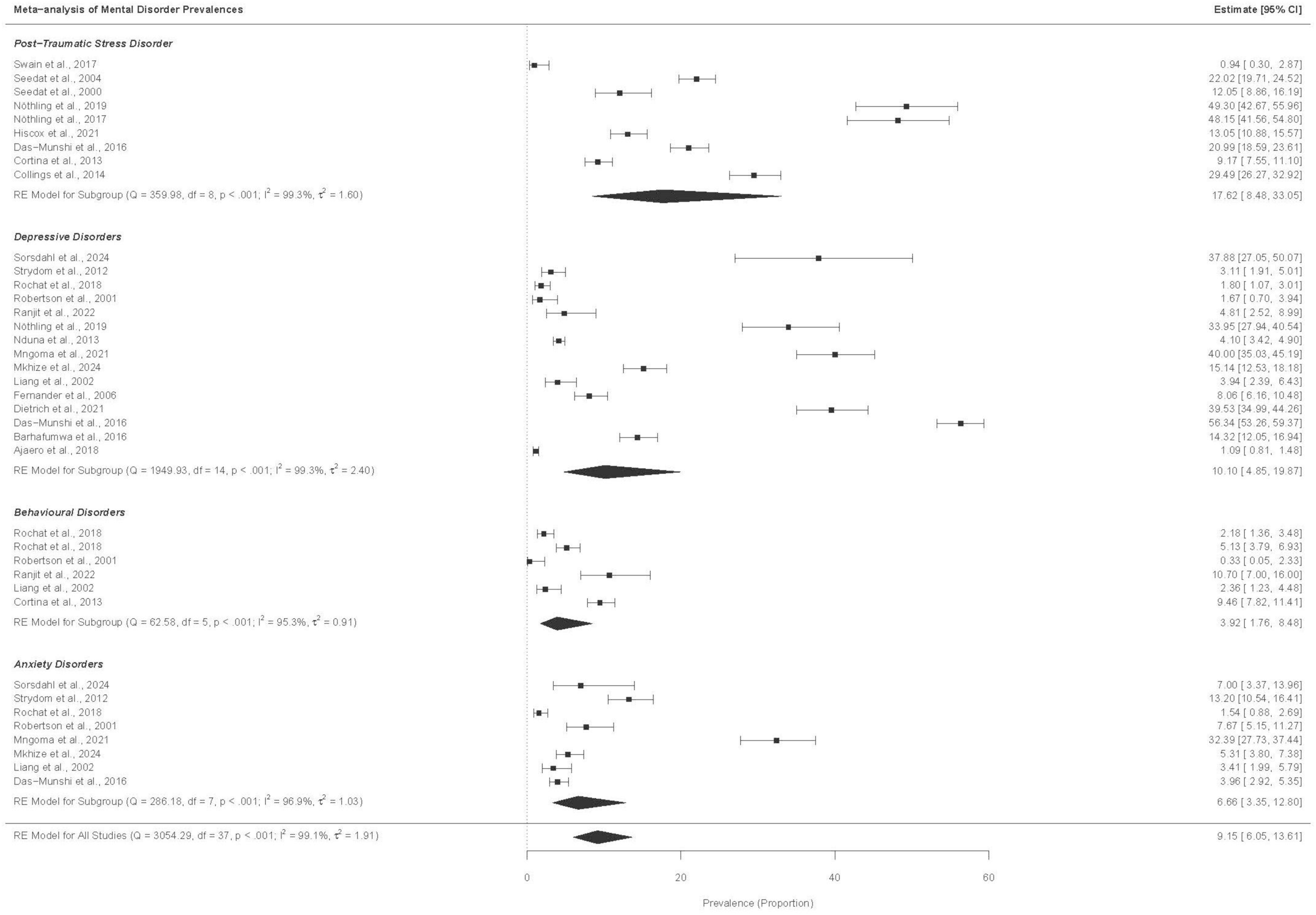
Forest plot of meta-analysis for prevalence of mental disorders with five or more studies.

The results of the univariate random effects meta-regression analysis to investigate potential sources of heterogeneity among included studies, using the various study characteristics as covariates, are presented in Supplementary Table S5. Prevalence rates in depression studies were significantly greater in papers published more recently (β=0.10 [0.00; 0.20], p=0.048). The prevalence of PTSD was significantly lower in studies that relied on symptom severity rather than diagnostic criteria to identify cases (β=-1.85 [-3.17; -0.53], p=0.006) and in studies that relied on observation for diagnosis (β=-3.44 [-5.52; -1.36], p=.001). For anxiety disorders, higher prevalence estimates were associated with increases in the mean age of the sample (β=0.32 [0.26; 0.37], p<0.0001) and with studies which assessed symptoms over a 1-week compared to 1-month recall period (β=2.00 [0.81; 3.19], p=0.001). Prevalence estimates for disruptive behaviour disorders were higher in school-compared to community-based surveys (β=1.40 [0.52; 2.27], p=0.002) but were lower in studies which assessed symptoms in the preceding 6-months compared to the preceding 1-month (β= -1.40 [-2.27; -0.62], p<0.002).

Results of the multivariate random effects meta-regression analysis are presented in Supplementary Table S6. For depressive disorders, prevalence estimates were higher in studies with school-rather than community-based samples (β=1.68 [0.52; 2.83], p=0.005) and lower in studies that used probability sampling (β=-1.36 [-2.72; -0.071, p<0.05). For PTSD prevalence estimates were lower in studies which used observations for diagnosis (β=-2.67 [-5.04; -0.30], p=0.027). For anxiety disorders higher prevalence estimates were associated with samples with a higher mean age (β=0.32 [0.26; 0.37], p<0.0001). No effects were observed for disruptive behaviour disorders.

### Suicidal thoughts and behaviours

Figure 3 illustrates the range of self-harm and suicidal phenomena reported. There were 30 prevalence estimates reported (see Supplementary Table S3 for full details of all studies). Prevalence estimates were reported for suicidal ideation (n= 10), suicide plan (n= 8), suicide intent (n=1), suicide attempt (n=9), completed suicide (n=1) and non-suicidal self-injury (n=1). Seven different tools were used to assess these outcomes, the most common of which were the *Youth Risk Behaviour Survey* (n=9), *Mini-International Neuropsychiatric Interview for Children and Adolescents (MINI-KID) suicidality scale* (n=6), and the *Suicidal Behaviour Questionnaire* (n=5).

The results of the meta-analysis for studies reporting suicidality are presented in Figure 5. Pooled prevalence estimates for suicidal ideation, plan and attempt were 12.0% [7.8; 18.0] (k=10, N=41489), 11.8% [7.7; 17.6] (k=8, N=39,928), and 10.3% [6.2; 16.6] (k=9, N=40,294), respectively. Very high heterogeneity was observed across the included studies for suicidal ideation (**I²**=99.63%, Q(9)=712.0, p<0.0001), suicide plans (I²=99.56%, Q(7)=572.3, p<0.0001), and suicide attempts (I²=99.69%, Q(8)=871.2, p<0.0001).

**Figure 5:**
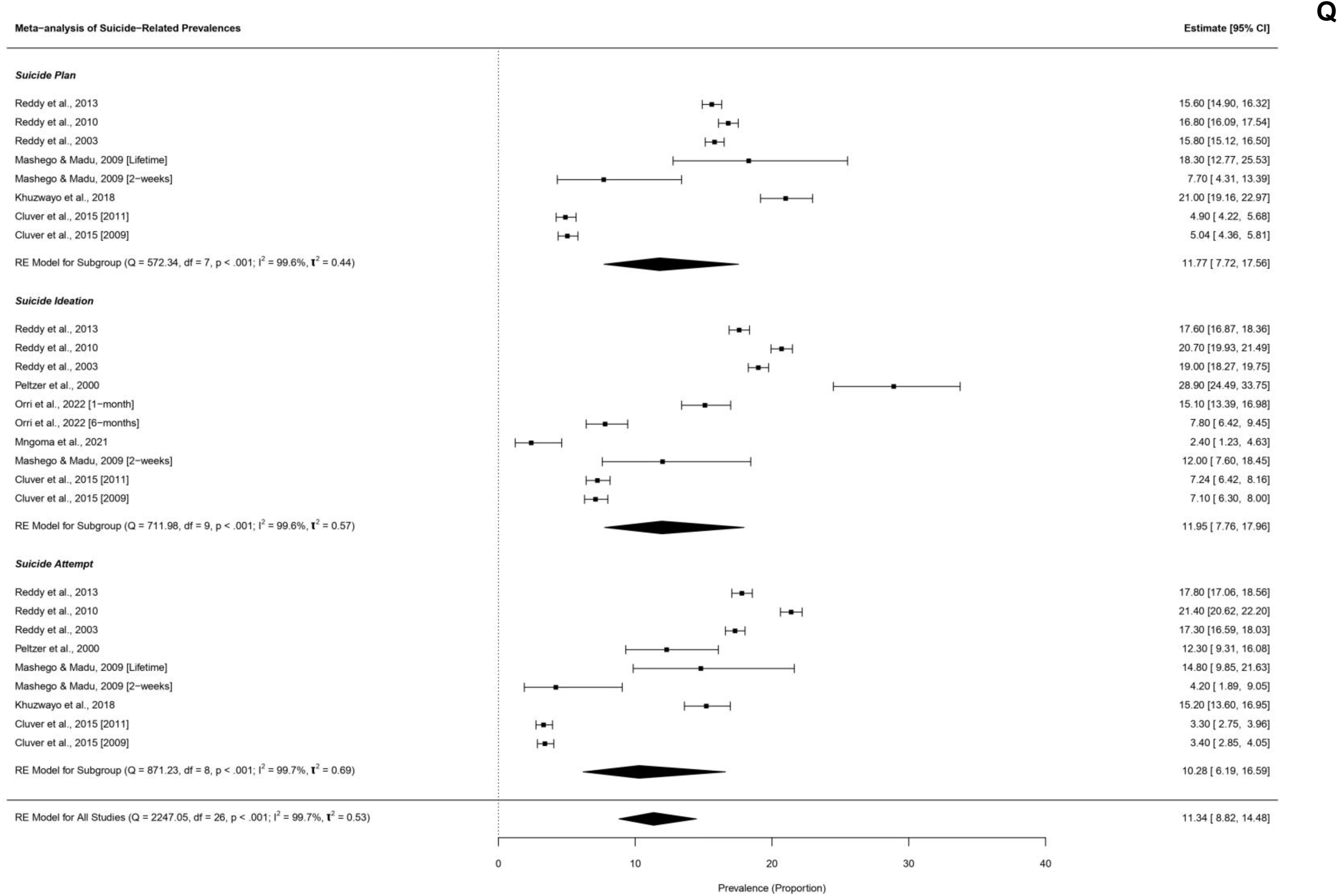
Forest plot of meta-analysis for prevalence of suicidal thoughts and behaviours

The results of the univariate random effects meta-regression analysis are presented in Supplementary Table S7. For suicidal ideation, prevalence rates were significantly lower in community-based surveys compared to school-based surveys (β= -1.10 [-1.71; -0.49], p=0.0004) and in studies published more recently (β=-0.07 [-0.12; -0.02], p = 0.004). Compared to studies that collected data in other provinces, rates of ideation were lower in the WC (β= -1.66 [-2.31;-1.02], p<0.0001), KZN (β=-2.81 [-3.80; -1.81], p<0.0001), Free State (β=-1.09 [-1.98;-0.21], p=0.016) and GP (β=-1.18 [-1.83; -0.53], p=0.0004), but were higher in the Eastern Cape (β=6.19 [4.16; 8.23], p<0.0001). Prevalence rates were also higher in studies that assessed lifetime prevalence as opposed to 1-month prevalence (β=1.38 [0.24; 2.53], p=0.018) but were lower in studies that assessed 1-week prevalence as opposed to 1-month prevalence (β=-1.42 [-2.73; -0.11], p=0.033). The prevalence of suicide plans was lower in community- vs school-based surveys (β=-1.34 [-1.62; -1.05], p<0.0001), the WC compared to other provinces (β=-1.32 [-1.51; -0.76], p<0.0001], longitudinal compared to cross-sectional studies (β=-1.34 [-1.62; -1.05], p<0.0001), and in studies which assessed 1-month prevalence compared to all other durations (p<0.0001). Prevalence estimates for suicide attempts were lower in community- vs school-based surveys (β=-1.66 [-2.13; -1.19], p<0.0001) and longitudinal vs cross-sectional studies (β= -1.66; [-2.13; -1.19], p<0.0001), but the prevalence was higher in samples which assessed symptoms over the period of 6-months (β = 0.15 [0.12; 0.19], p<0.0001), 1-year (β=0.12 [0.08; 0.16], p<0.0001), and lifetime (β=0.10 [0.07; 0.14], p<0.0001) comparted to 1-month.

The results of the multivariate random effects meta-regression analysis are presented in Supplementary Table S8. Compared to data collected in other provinces, the prevalence of suicidal ideation was lower in the Western Cape (β=-0.88 [-1.23; -0.54], p<0.0001), KwaZulu-Natal (β=-1.99 [-2.73; -1.24], p<0.0001), and the Free State (β=-1.03 [-1.69; -0.36], p=0.003) but higher in the Eastern Cape (β=4.16 [2.95; 5.36], p<0.0001). Counter-intuitively, prevalence estimates were lower in studies which assessed ideation in the past 6-months compared to the past month (β=-0.74 [-1.11; -0.37], p=0.0001). For suicide plans, prevalence estimates were lower in school- vs community-based studies (β=-1.19 [-1.70; -0.68], p<0.0001) and in studies which assessed 2-week vs 1-month prevalence (β=-0.99 [-1.74; -0.23], p=0.010. For geographic location, prevalence estimates were lower in the WC (β=-0.38 [-0.62; -0.15], p=0.001). For suicide attempts, prevalence estimates were lower in school- vs community-based studies (β=-1.44 [-2.45; -0.42], p=0.006) and in studies which assessed attempts in the past 2-weeks vs 1-month (β=-1.38 [-2.38; -0.38], p =0.007).

### Quality assessment

The results of the quality assessment are presented as supplementary materials (see Table S2 and S3). All included studies were assessed as being of sufficient quality indicating that they all met the essential criteria to be considered reliable and useful. Nonetheless the confidence intervals in some studies were large indicating that some prevalence estimates might not be reliable because of small sample sizes. Several studies did not report information about the study design and data collection (evidenced from the missing data in Tables S2 and S3).

## DISCUSSION

This systematic review of the mental health of children and adolescents in SA, identified 40 studies with 56 prevalence estimates for mental disorders and 30 prevalence estimates for suicidality. Random effects meta-analysis suggests a pooled prevalence for all disorders of 14.9% [9.9; 19.9], across all recall periods. The prevalence estimate is broadly aligned with the results of a meta-analysis reporting a worldwide prevalence of 13.4% [11.3–15.9] for mental disorders in children and adolescents[18] and the 2019 *Global Burden of Disease* estimate of 11.6% among 5- to 24-year-olds, globally[34]. Our meta-analysis also yielded pooled prevalence estimates for depression (10.1% [4.9; 19.9]), anxiety (6.7% [3.4; 12.8]), PTSD (17.6% [8.5; 33.1]), and disruptive behaviour disorders (3.9% [1.8; 8.5]). For all other disorders, there were either no studies or too few studies for meta-analysis. Crucially, the available evidence on the prevalence of mental disorders is significantly heterogenous, making it difficult to say anything reliable about the burden of mental disorders among children and adolescents in SA.

Prevalence estimates we observed for depression (10.1%) and anxiety disorders (6.7%) are broadly aligned with global estimates in studies which assess symptom severity[35,36]. Worldwide, studies which use strict diagnostic criteria and structured interviews to identify cases[37], report pooled prevalence rates of 2.6% [1.7; 3.9] for depression and 6.5% [4.7; 9.1] for anxiety disorders[37]. It is remarkable that we observed a pooled prevalence of 17.6% for PTSD, which is probably a consequence of high rates of exposure to violence[13] and sexual assault[14] among SA’s youth. Even though this is probably an over-estimate of the true prevalence of PTSD, it does highlight the likelihood of a substantial need for interventions to address trauma among SA’s youth. It is important that future studies establish accurate prevalence estimates for trauma-related disorders, so that adequate provision can be made for services. This is particularly important given the persistence of trauma-related disorders and their association with other serious mental health problems, like substance use and severe depressive illness, when left untreated[38].

Research on child and adolescent mental health in SA has disproportionately focused on common disorders like depression, anxiety and PTSD, with complete neglect of other debilitating disorders such as autism spectrum disorder, intellectual disability, psychotic illnesses, learning disorders, substance use disorders, and obsessive-compulsive disorders. Importantly, we did not identify any studies reporting on cognitive impairment, mental well-being, quality of life, mental health literacy, or mental health awareness, that met inclusion criteria for this review, demonstrating a clear imbalance towards studies that conceptualize child and adolescent mental health through the lens of psychopathology rather than wellness. Moreover, little attention has been paid to assessing the level of functioning and role impairment associated with mental disorders among SA’s youth.

Even though suicide is one of the leading causes of death among young people globally[39], there are only 11 studies reporting suicidal behaviours and self-harm among SA youth. As with mental disorders, these studies are remarkably heterogenous and provide an incomplete and contradictory picture of suicide risk among the country’s youth. Most of the data in this area come from 3 national youth risk surveys which report data that is contradictory. In the three national youth risk surveys[40–42] the 6-month prevalence of suicide attempts is reported as 17.3% (2002), 21.4% (2008) and 17.8% (2011), suggesting that almost 1 in 5 children has attempted suicide in the past 6 months, which is implausible. A recent systematic review and meta-analysis reported that globally the lifetime prevalence of suicide attempts among youth was 7.0% [6.0-8.1%][43], which is significantly lower than the rates reported in the *South African Youth Risk Behaviour Surveys*. Furthermore, the prevalence of suicide attempts reported in the national youth risk surveys is consistently higher than the prevalence reported for suicide plan, which is also surprising given that formulating a suicide plan is a prerequisite for attempting suicide. Considerably more work needs to be done to understand the epidemiology of suicidal behaviour among children and adolescents in SA. However, such work is challenging for several reasons including that suicide is still highly stigmatised in SA and questions about suicidal thoughts and behaviours are not easy to translate into indigenous languages. More work is needed to discover how to ask young people about suicidal phenomena in ways that are both culturally appropriate and comprehensive so that the data collected is meaningful [44].

There are several methodological concerns with the studies we identified. Studies on child and adolescent mental health in SA use self-report screening instruments which assess symptom severity rather than diagnostic criteria. Furthermore, many studies used instruments which had not been validated and/or culturally adapted. These findings about the methodological shortcomings of available studies are highly consistent with the results of similar reviews conducted by members of our team in Greece[16] and Brazil[17]. The geographic distribution of studies is very uneven with most studies conducted in the WC, KZN and GP, the three provinces with the largest populations in the country. The WC and GP are also the provinces with the highest per capita GDP, which may partially explain the preponderance of studies in these regions. Taken together, this makes it difficult to say anything meaningful about the prevalence of child and adolescent mental health in SA. The results of this systematic review highlight the need for a robust epidemiological survey with a nationally representative sample, using well validated instruments to investigate a wide range of mental disorders.

### Strengths and limitations

This is the first systematic review of prevalence studies on child and adolescent mental health in SA. The methods used are rigorous and include a wide range of search terms to identify studies that focus on all aspects of mental health, including both pathology and wellness, and clear and relevant inclusion criteria have been applied to identify studies that describe the mental health burden in children and adolescents. The conclusions we have drawn are, however, limited by the small number of studies that have been done in this area and the profound heterogeneity across included studies. We also excluded clinical samples and treatment seeking samples, and we did not report treatment rates, all of which are important indicators for allocating mental health resources. There is also an awareness that studies originating in the global south do face disproportionate publication biases and acceptances in international journals which may have contributed to limited studies that were available for inclusion[45].

## CONCLUSION

While studies suggest high rates of disorders among children and adolescents in SA, we cannot firmly establish nationwide estimates to inform advocacy and policy. There is an urgent need for a national survey of child and adolescent mental health, using well validated instruments that can assess a wide range of disorders among a representative sample of young people. This will provide the essential platform for guiding the allocation of scarce mental health resources to support and protect the mental health of SA youth. In the absence of such a survey there is no empirical basis for making data driven decisions on child and adolescent mental health to plan programs and services and set priorities.

## Data Availability

All data produced in the present work are contained in the manuscript and supplementary materials.

## Acknowledgements

None.

## Contributors

GAS and LEM: study design, data analysis, data interpretation and contributing text to the manuscript.

JB: data interpretation and drafting of manuscript.

CBS: overseeing data extraction, data interpretation and contributing text to the manuscript.

DJ data management, statistical analysis, interpretation of findings and review of the manuscript.

NM, YC, NH and MM: data extraction, preparation of tables, manuscript formatting, and quality checking.

TCP: data management and data quality checking.

SR: input on data extraction, data interpretation and contributing to the manuscript.

DJS, SS, MT, SS, AL, IP, BC, SP, RN, MY, XH, EC, ECM, ZM: data interpretation and contributing text to the manuscript.

## Funding

This work was funded by the Stavros Niarchos Foundation (SNF) *Global Center for Child and Adolescent Mental Health* at the *Child Mind Institute*. Additional support was provided by the South African Medical Research Council.

## Competing Interests

None to declare.

## Patient consent

Not required

## Notes

### Competing Interest Statement

The authors have declared no competing interest.

## References

1 Benton TD, Beers L, Carlson G, et al. The Declaration of the National Emergency in Child and Adolescent Mental Health: It Takes a Village. Child Adolesc Psychiatr Clin N Am. 2024;33:277–91. doi: 10.1016/j.chc.2024.03.001

2 McGorry PD, Mei C, Dalal N, et al. The Lancet Psychiatry Commission on youth mental health. Lancet Psychiatry. 2024;11:731–74. doi: 10.1016/S2215-0366(24)00163-9

3 Piao J, Huang Y, Han C, et al. Alarming changes in the global burden of mental disorders in children and adolescents from 1990 to 2019: a systematic analysis for the Global Burden of Disease study. Eur Child Adolesc Psychiatry. 2022;31:1827–45. doi: 10.1007/S00787-022-02040-4/FIGURES/5

4 Kessler RC, Angermeyer M, Anthony JC, et al. Lifetime prevalence and age-of-onset distributions of mental disorders in the World Health Organization’s World Mental Health Survey Initiative. World Psychiatry. 2007;6:168.

5 Kim-Cohen J, Caspi A, Moffitt TE, et al. Prior Juvenile Diagnoses in Adults With Mental Disorder: Developmental Follow-Back of a Prospective-Longitudinal Cohort. Arch Gen Psychiatry. 2003;60:709–17. doi: 10.1001/ARCHPSYC.60.7.709

6 Kieling C, Baker-Henningham H, Belfer M, et al. Child and adolescent mental health worldwide: Evidence for action. The Lancet. 2011;378:1515–25. doi: 10.1016/S0140-6736(11)60827-1/ATTACHMENT/2BE1DC68-0B55-4815-8BC8-CAB4A84B1071/MMC1.PDF

7 Costello EJ, Burns BJ, Angold A, et al. How Can Epidemiology Improve Mental Health Services for Children and Adolescents? J Am Acad Child Adolesc Psychiatry. 1993;32:1106–17. doi: 10.1097/00004583-199311000-00002

8 Ford T. Practitioner Review: How can epidemiology help us plan and deliver effective child and adolescent mental health services? Journal of Child Psychology and Psychiatry. 2008;49:900–14. doi: 10.1111/J.1469-7610.2008.01927.X

9 Casella CB, Kousoulis AA, Kohrt BA, et al. Data gaps in prevalence rates of mental health conditions around the world: a retrospective analysis of nationally representative data. Lancet Glob Health. 2025;13:e879–87. doi: 10.1016/S2214-109X(24)00563-1

10 Erskine HE, Baxter AJ, Patton G, et al. The global coverage of prevalence data for mental disorders in children and adolescents. Epidemiol Psychiatr Sci. 2017;26:395–402. doi: 10.1017/S2045796015001158

11 Jorns-Presentati A, Napp AK, Dessauvagie AS, et al. The prevalence of mental health problems in sub-Saharan adolescents: A systematic review. PLoS One. 2021;16:e0251689. doi: 10.1371/JOURNAL.PONE.0251689

12 Hall K. Child Poverty. Statistics of Children in South Africa. 2024;7. http://childrencount.uct.ac.za/indicator.php?domain=2&indicator=98#:~:text=Using%20Stats%20SA%27s%20lower%20bound,were%20not%20getting%20enough%20nutrition. (accessed 23 January 2025)

13 Richter LM, Mathews S, Kagura J, et al. A longitudinal perspective on violence in the lives of South African children from the Birth to Twenty Plus cohort study in Johannesburg-Soweto. South African Medical Journal. 2018;108:181–6. doi: 10.7196/SAMJ.2018.v108i3.12661

14 Seedat M, Van Niekerk A, Jewkes R, et al. Violence and injuries in South Africa: prioritising an agenda for prevention. The Lancet. 2009;374:1011–22. doi: 10.1016/S0140-6736(09)60948-X/ASSET/64E3FA8F-2EC8-4938-B2A2-866534036DE2/MAIN.ASSETS/GR1.JPG

15 Mumbauer AE, Stein DJ, Wolvaardt GG. Targeting youth mental health in a demographically young country: a scoping review focused on South Africa. International Review of Psychiatry. Published Online First: 29 November 2024. doi: 10.1080/09540261.2024.2429648

16 Koumoula A, Marchionatti LE, Caye A, et al. The science of child and adolescent mental health in Greece: a nationwide systematic review. Eur Child Adolesc Psychiatry. 2023;33:3359–75. doi: 10.1007/s00787-023-02213-9

17 Marchionatti LE, Campello AC, Veronesi JA, et al. The science of child and adolescent mental health in Brazil: a nationwide systematic review and compendium of evidence-based resources. 2024.

18 Polanczyk G V., Salum GA, Sugaya LS, et al. Annual Research Review: A meta-analysis of the worldwide prevalence of mental disorders in children and adolescents. Journal of Child Psychology and Psychiatry. 2015;56:345–65. doi: 10.1111/jcpp.12381

19 Brooke-Sumner C, BJ, , RS, MZ, ML, SG. Systematic review of prevalence of mental disoders in children and adolescents in South Africa. PROSPERO. 2024.

20 Thomas J, BJ, & GS. EPPI-Reviewer 4.0: software for research synthesis. EPPI-Centre Software. . London, UK: Social Science Research Unit, Institute of Education 2010.

21 Ouzzani M, Hammady H, Fedorowicz Z, et al. Rayyan-a web and mobile app for systematic reviews. Syst Rev. 2016;5:1–10. doi: 10.1186/S13643-016-0384-4/FIGURES/6

22 Hoy D, Brooks P, Woolf A, et al. Assessing risk of bias in prevalence studies: modification of an existing tool and evidence of interrater agreement. J Clin Epidemiol. 2012;65:934–9. doi: 10.1016/j.jclinepi.2011.11.014

23 Munn Z, Moola S, Riitano D, et al. The Development of a Critical Appraisal Tool for Use in Systematic Reviews: Addressing Questions of Prevalence. Int J Health Policy Manag. 2014;3:123–8. doi: 10.15171/IJHPM.2014.71

24 Branscum AJ, Gardner IA, Johnson WO. Estimation of diagnostic-test sensitivity and specificity through Bayesian modeling. Prev Vet Med. 2005;68:145–63. doi: 10.1016/J.PREVETMED.2004.12.005

25 Rogan WJ, Gladen B. ESTIMATING PREVALENCE FROM THE RESULTS OF A SCREENING TEST. Am J Epidemiol. 1978;107:71–6. doi: 10.1093/OXFORDJOURNALS.AJE.A112510

26 Devleesschauwer B, Torgerson P, Charlier J, et al. Tools for Prevalence Assessment Studies [R package prevalence version 0.4.1]. *CRAN: Contributed Packages*. Published Online First: 3 June 2022. doi: 10.32614/CRAN.PACKAGE.PREVALENCE

27 Speybroeck N, Devleesschauwer B, Joseph L, et al. Misclassification errors in prevalence estimation: Bayesian handling with care. Int J Public Health. 2013;58:791–5. doi: 10.1007/S00038-012-0439-9/TABLES/4

28 R: The R Project for Statistical Computing. 2025. https://www.r-project.org/ (accessed 10 April 2025)

29 Viechtbauer W. Conducting Meta-Analyses in R with the metafor Package. J Stat Softw. 2010;36:1–48. doi: 10.18637/JSS.V036.I03

30 Page MJ, McKenzie JE, Bossuyt PM, et al. The PRISMA 2020 statement: an updated guideline for reporting systematic reviews. BMJ. 2021;372. doi: 10.1136/BMJ.N71

31 Whiting P, Savović J, Higgins JPT, et al. ROBIS: A new tool to assess risk of bias in systematic reviews was developed. J Clin Epidemiol. 2016;69:225. doi: 10.1016/J.JCLINEPI.2015.06.005

32 Burrows S, Laflamme L. Suicide among urban South African adolescents. Int J Adolesc Med Health. 2008;20:519–28. doi: 10.1515/IJAMH.2008.20.4.519/PDF/FIRSTPAGE

33 Hall K. Demography of South Africa’s children . Cape Town 2022.

34 Kieling C, Buchweitz C, Caye A, et al. Worldwide Prevalence and Disability From Mental Disorders Across Childhood and Adolescence: Evidence From the Global Burden of Disease Study. JAMA Psychiatry. 2024;81:347–56. doi: 10.1001/JAMAPSYCHIATRY.2023.5051

35 Shorey S, Ng ED, Wong CHJ. Global prevalence of depression and elevated depressive symptoms among adolescents: A systematic review and meta-analysis. British Journal of Clinical Psychology. 2022;61:287–305. doi: 10.1111/BJC.12333

36 Lu B, Lin L, Su X. Global burden of depression or depressive symptoms in children and adolescents: A systematic review and meta-analysis. J Affect Disord. 2024;354:553–62. doi: 10.1016/J.JAD.2024.03.074

37 Polanczyk G V., Salum GA, Sugaya LS, et al. Annual Research Review: A meta-analysis of the worldwide prevalence of mental disorders in children and adolescents. Journal of Child Psychology and Psychiatry. 2015;56:345–65. doi: 10.1111/JCPP.12381

38 Lockwood E, Forbes D, Lockwood E, et al. Posttraumatic Stress Disorder and Comorbidity: Untangling the Gordian Knot. Psychological Injury and Law 2014 7:2. 2014;7:108–21. doi: 10.1007/S12207-014-9189-8

39 World Health Organisation. Suicide worldwide in 2019 Global Health Estimates. Geneva 2021.

40 Reddy S, Panday S, Swart D, et al. Umthente Uhlaba Usamila Umthente Uhlaba Usamila Umthente Uhlaba Usamila REPORT PREPARED FOR THE SOUTH AFRICAN NATIONAL DEPARTMENT OF HEALTH. Cape Town 2003.

41 Reddy S, James S., Sewpaul R., et al. Umthente Uhlaba Usamila Umthente Uhlaba Usamila The 2 nd South African National Youth Risk Behaviour Survey 2008 The 2 nd South African National Youth Risk Behaviour Survey 2008. 2010.

42 Reddy SP, James S, Sewpaul R, et al. Umthente Uhlaba Usamila: the 3rd South African National Youth Risk Behaviour Survey 2011. South African Medical Research Council 2015.

43 Van Meter AR, Knowles EA, Mintz EH. Systematic Review and Meta-analysis: International Prevalence of Suicidal Ideation and Attempt in Youth. J Am Acad Child Adolesc Psychiatry. 2023;62:973–86. doi: 10.1016/J.JAAC.2022.07.867

44 Bantjes J, Breet E, Kagee A, et al. Suicide prevention in sub-Saharan Afrrica: Cultural considerations and opportunities to advance suicide research and practice. In: Schouler-Ocak M, Khan M, eds. Suicide Across Cultures: . Oxford, UK: Oxford University Press 2024:184–204.

45 Draper CE, Barnett LM, Cook CJ, et al. Publishing child development research from around the world: An unfair playing field resulting in most of the world’s child population under-represented in research. Infant Child Dev. 2023;32:e2375. doi: 10.1002/ICD.2375

